# Evaluation of HIV Viral Load Surveillance System, National Perspective in Tanzania

**DOI:** 10.1101/2024.01.17.24301295

**Authors:** Peter Richard Torokaa, Loveness John Urio, Ambwene Mwakalobo, Alex Sifaeli Magesa, James Allan, David J. Osima, Focus Medard Shao, Joseph Mziray, Yulitha Barnabas, Agricola Joachim, Mtebe Majigo

## Abstract

**Background:** Human immunodeficiency virus (HIV) infection is still a global public health problem. The number of HIV-positive individuals was estimated to be 38.0 million worldwide in 2020, with 2.78 million being children and adolescents aged 0 to 19 years. We aimed to assess the usefulness and system attributes of the HIV Viral Load (HVL) surveillance system in Tanzania and determine whether it meets its objectives.

**Methodology:** This was a descriptive cross-sectional analysis conducted to describe the attributes and usefulness of the HVL surveillance system. From January through December 2022, we consistently captured HVL data, which we extracted and analysed. The system description, case definition, and eligibility criteria were obtained from the system guidelines. We used Updated Guidelines for Evaluating Public Health Surveillance Systems from the Centre for Disease Control and Prevention (CDC) to evaluate system attributes. We observed the data entry to determine the simplicity and time spent entering data into the system. Turnaround time were determined from the sample collected at the spoke to the result returned to the clients.

**Results:** The HVL surveillance system is implemented by 90.6% (4314/4764) of facilities in the country. The overall national data completeness was 97.4%. The system’s sensitivity was 84.4%, with a predictive value positive (PVP) of 52% and a national data accuracy of 97%. Of eight data clerks interviewed, 100% (8/8) said the system was simple to use.

**Conclusion:** The system’s overall performance achieved its goals was satisfactory. The system was representable, simple, flexible, and sensitive. Turnaround time was at a median of 13 days.

## Introduction

Human immunodeficiency virus (HIV) infection is still a global public health problem. The number of HIV-positive individuals was estimated to be 38.0 million worldwide in 2020, with 2.78 million being children and adolescents aged 0 to 19 years. In the same year, 120,000 young people under 18 years died due to Acquired immunodeficiency syndrome (AIDS)-related causes, while 310,000 young people under 18 years contracted HIV for the first time (1). The target number of 370,000 new HIV infections for 2025 will triple to 1.2 million persons if the current trends continue (2). The data indicate the need for countries to monitor HIV prevention, care, and treatment interventions effectively.

The Joint United Nations Programme on HIV/AIDS established the 95-95-95 targets, meaning 95% of all people living with HIV should know their HIV status, 95% of all people with diagnosed HIV infection should receive sustained antiretroviral therapy (ART) and 95% of all people receiving ART to have viral suppression by 2030 (3). Sixteen nations, eight sub-Saharan African nations, are nearing the 95-95-95 targets, with Botswana, Eswatini, Rwanda, Tanzania, and Zimbabwe already achieving this (3). HIV viral load (HVL) monitoring is the gold standard for identifying treatment failure. Its accessibility in resource-limited settings has been severely constrained due to high cost (40–85 USD/test), complicated specimen collection procedures, and transport requirements (4). HVL testing and monitoring has complex technical and logistic requirements requiring continuous expert guidance and a reliable quality management system. Some countries, including Tanzania, have adopted a hub-and-spoke model coupled with clustering clinics and efficient specimen collection, transportation and referral systems (5).

World health organization (WHO) advises that testing be made available to everyone at risk for HIV. In-depth and efficient HIV prevention, testing, and treatment services should be carried out for those at higher risk of contracting the virus (6). HVL testing for monitoring HIV care and treatment among people living with HIV was started in Tanzania in 2017, with the growth of the hub-and-spokes approach for sample processing and transportation and the scaling up of testing facilities. Tanzania has seen a notable rise in HVL testing since 2017 (5). The nation has concentrated on tracking the quantity of HIV clients who are tested and following up with them along the HVL cascade, especially in cases when the client has a high viral load. To guarantee efficient HIV treatment, the nation’s actions are essential (7). The objectives of HVL surveillance system are tracking HVL testing, specimen, and results, regularly monitor and evaluate the efficiency of specimen transportation, processing, testing, timely delivery of HVL test results turnaround time (TAT), provide information on antiretroviral (ARV) drug efficacy (viral load suppression), provide indication on patient adherence to treatment regimen, determine trends on ART failure and determine the magnitude of drug resistance.

It is essential to put in place a tracking system for HVL test, specimens, and results and to routinely assess how well specimen transportation, processing, testing, and delivery are crucial. Resources must be allocated to the surveillance and evaluation of the HVL systems to ensure the system’s usefulness and objectives are met. Since the start of HVL surveillance system in 2017, it has not been evaluated to check its performance. We aimed to assess the usefulness of the HIV viral load surveillance system in Tanzania and determine whether it meets its objectives.

## Material and Methodology

### Design and settings

This was a descriptive cross-sectional analysis to describe the attributes of the HVL surveillance system. We consistently captured HVL data collected from January through December 2022. The HVL data are hosted in the Ministry of Health at the National AIDS Control Program in the Open Laboratory Data Repository (OpenLDR) database. The database contains all HVL data from hubs, spokes, and testing laboratories in the country. OpenLDR is the national database that collects all testing information data from other laboratory information systems for dashboard visualization. The data in the database are available from all twenty (20) conventional laboratories.

We purposefully selected Dodoma Regional Referral Hospital (DRRH) and Makole Health Centre (MHC) for the data verification due to the high volume of clients. DRRH serves the country’s capital city in Tanzania and is one of the HVL laboratories. DRRH is the central zone for HVL testing, covering Singida and Dodoma regions. MHC is the health centre that serves as the council hospital at Dodoma City Council, with a population of 765,179, according to the 2022 census.

### Population under surveillance

The surveillance system included all clients living with HIV who started ART treatment and were eligible for viral load testing, where every client six or twelve months after starting ART and then every 12 months (yearly), and any patients with clinical or suspected immunological failure. We extracted data like the information, such as age, gender, and HVL results.

### HVL surveillance system data flow

The HVL surveillance system collects samples of care and treatment centres (CTC) (spokes). HVL testing adheres to the pre-examination, examination, and post-examination phases typical of clinical laboratory testing methods. Samples from spokes are sent to hub, where eligible samples are separated into plasma after cross-checking. After that, samples are taken to a PCR laboratory for analysis. The results are then transmitted via hub to healthcare facilities. Results from the hubs are input into the CTC2 database and then into the CTC3 national database. Data is then sent to the OpenLDR database for HVL surveillance visualization. Fig 1 depicts the information flow from one level to the next. At the national level, the dashboard analyses data and visualizes it.

**Fig 1.**
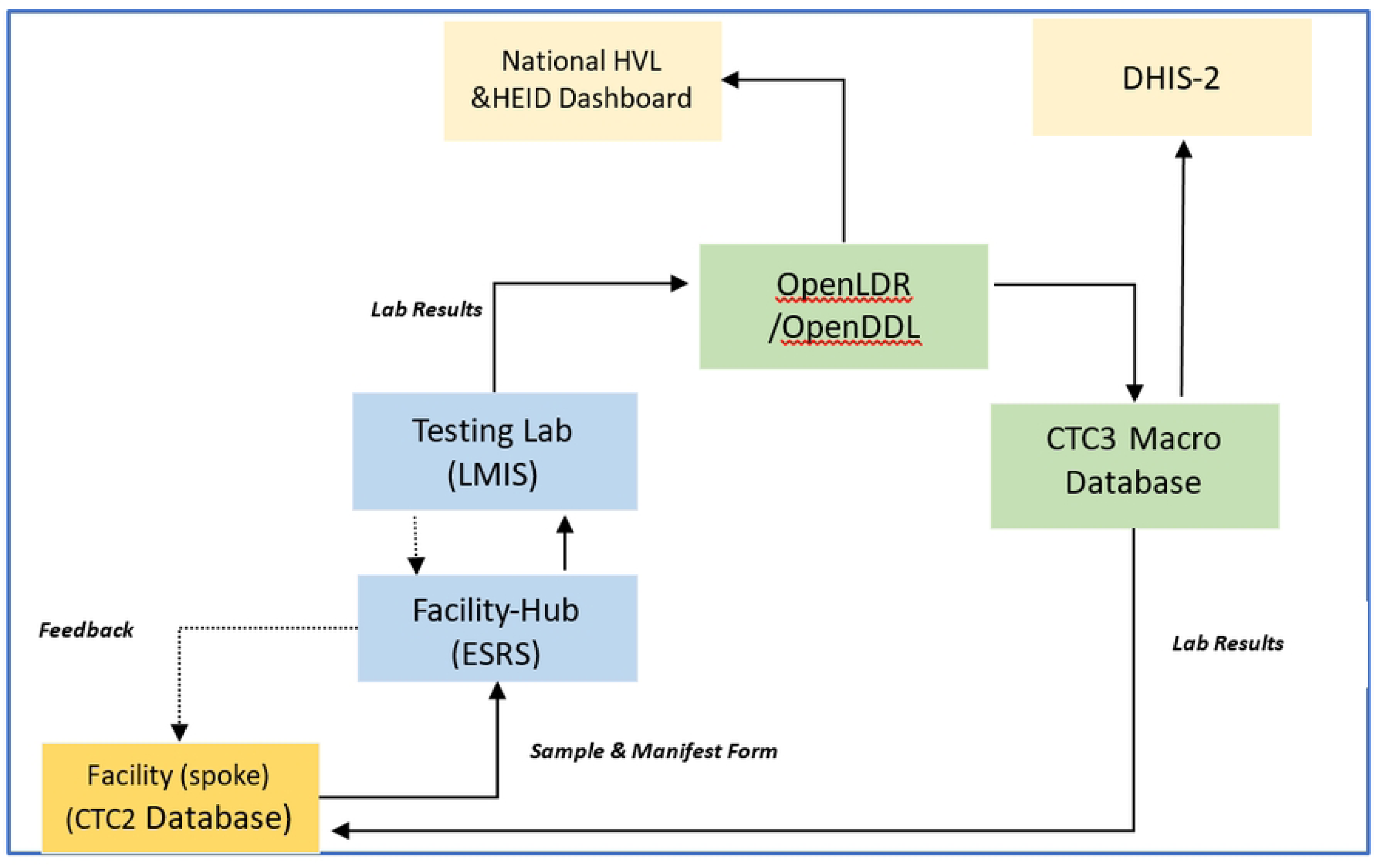
Illustrates the electronic laboratory data flow. Source from NACP National HIV Viral Load, HIV Early Infant Diagnosis and HIV Drug Resistance Testing Guidelines. ***HVL****- HIV Viral Load, **HEID**- HIV Early Infant Diagnosis, **OpenLDR**- Open Laboratory Data Repository, **DHIS2**- District Health Information System, **LMIS**- Laboratory Management Information System, **eSRS**- Electronic Sample Referral System, **CTC**- Centre for Treatment and Care*.

### Data collection procedure

Data from the national level for all twenty laboratories were extracted from OpenLDR on 27^th^ February 2023 and exported into Microsoft Excel. We extracted variables like gender, age, testing laboratory zones, sample received date, results authorized date, and reason for testing. We selected two health facilities for data verification from the laboratory information systems and that of the CTC2 database (Dodoma Regional Referral Hospital and Makole Health Centre). Data verified was the clients’ results entry from the Laboratory information system and the CTC2 database. Using a structured interview guide, we interviewed laboratory and CTC2 database data clerks and CTC in charge. The system attributes assessed by the interview were simplicity, acceptability, flexibility, and system usefulness.

### Definition of Surveillance system attributes

***Usefulness:*** Describes how a public health surveillance system contributes to preventing and controlling adverse health-related events, including an improved understanding of the public health implications of such events. A public health surveillance system can also be useful if it helps to determine that an adverse health-related event previously thought to be unimportant is important. In addition, data from a surveillance system can contribute to performance measures, including health indicators used in needs assessments and accountability systems. We assessed the usefulness of the surveillance system by interviewing the CTC in-charges on the use of information generated by the system, how the information used to improve the programs and make the proper decisions.

***Simplicity***: Refers to both its structure and ease of operation. Surveillance systems should be as simple as possible while still meeting their objectives. We assessed this attribute by interviewing and observing the data entry in the system.

***Flexibility*:** This is the ability of the surveillance system to adapt to changing information needs or operating conditions with little additional time, personnel, or allocated funds. Flexible systems can accommodate new health-related events, changes in case definitions or technology, and variations in funding or reporting sources. In addition, systems that use standard data formats (e.g., in electronic data interchange) can be easily integrated with other systems and thus might be considered flexible. We assessed the flexibility of the surveillance system to integrate with other systems and the addition of the other surveillance system without adding time, personnel, and funds.

***Data Quality:*** Refers to data quality and reflects the completeness and validity of the data recorded in the public health surveillance system. We assessed the completeness of data and validity by evaluating the proportion of completeness for the data sent to the national level and data correctness. Furthermore, we evaluated data consistency by comparing the data from the laboratory information system and that in the CTC2 database.

***Acceptability:*** Acceptability reflects the willingness of persons and organizations to participate in the surveillance system. In this attribute, we assessed the willingness to participate in the surveillance system by interviewing the laboratory data clerks, CTC2 database data clerks, and CTC in charge.

***Sensitivity***: The sensitivity of a surveillance system can be considered on two levels. First, at the level of case reporting, sensitivity refers to the proportion of cases of a disease (or other health-related event) detected by the surveillance system. Second, sensitivity can refer to the ability to detect outbreaks and monitor changes in the number of cases over time. We assessed the number of eligible clients to be taken HVL samples and those collected during the study period.

***Predictive Value Positive:*** Predictive value positive (PVP) is the proportion of reported cases with a health-related event under surveillance. We assessed the PPV by evaluating the correct eligible clients tested for HVL to total clients tested for HVL.

***Representativeness:*** A representative public health surveillance system accurately describes the occurrence of a health-related event over time and its distribution in the population by place and person. We assessed the distribution of cases in the country and the sites (number of involved active health facilities) where the cases were identified.

***Timeliness***: Reflects the speed between steps in a public health surveillance system. We assessed the Time spent from sample collection to laboratory report to the requesting facility.

***Stability***: Refers to the reliability (i.e., the ability to collect, manage, and provide data properly without failure) and availability (the ability to be operational when needed) of the public health surveillance system. We assessed the number of events of the system outage from laboratory information systems and that of the CTC2 database. We interviewed data clerks to provide their insight into the system’s stability.

### Data processing and analysis

We evaluated the surveillance system’s attributes using CDC guidelines for evaluating public health surveillance systems (8). The system’s attributes evaluated include usefulness, simplicity, flexibility, representativeness, timeliness, data quality, acceptability, sensitivity, predictive value positive, and stability of the HIV viral load system. Data were entered, cleaned, and stored in Microsoft Excel, and the system attributes were analysed using Microsoft Excel and Stata version 15.1. We calculated the frequency and proportions to evaluate the system attributes.

## Ethical considerations

The permission to conduct this study was obtained from the Ministry of Health through Ref. no. PA.160/291/01A/ 41 and data verification access was obtained from the President’s Office Regional Administration and Local Government through permission Ref. no. AB.81/228/01.

## Results

### Participants Characteristics

We extracted data at the national level for 1,048,575 HVL samples collected from January to December 2022. Females numbered 720,228 (68.7%), while 20,466 (2%) were missing gender information. The participants aged above 30 years old were 820,472 (78.3%). Lake zone had a high number of samples tested, 274,728 (26.2%), and the least zone was the Southern zone, 44,434 (4.2%) (Table 1). All eight data clerks (100%) interviewed provided their insight that the system was useful for providing real-time information about HIV clients.

**Table 1.**
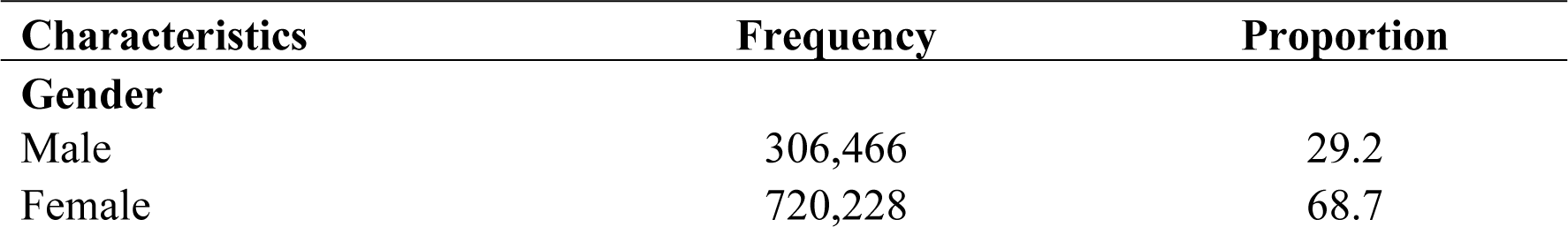
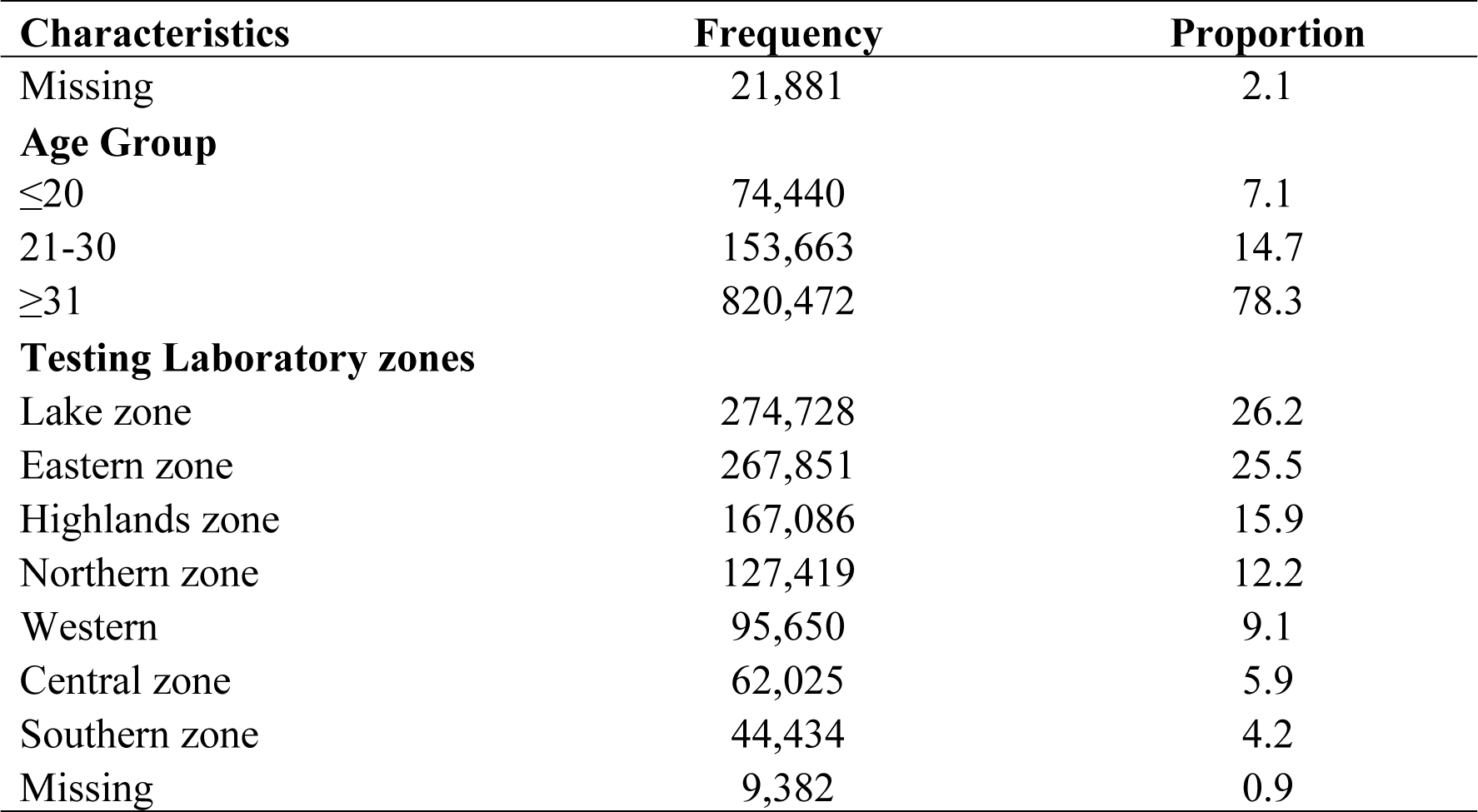
Demographic characteristics of participants (N= 1,048,575)

### HIV viral load surveillance system attributes

#### Usefulness

The two CTC in charges interviewed regarding the HIV viral load surveillance system usefulness both acknowledged that the system was useful in the context of easy availability of the information. The system was able to produce different summary reports that are used for the facility decision making and the entire program decision. The system was able to generate the cross sectional and cohort reports which shows like the trends of client’s enrolment, eligible clients for HVL testing and viral load suppression summary.

#### Simplicity

All four laboratory data clerks (100%) interviewed reported that both Tillehub and Electronic Viral Load Information Management System (EVLIMS) systems are easy to use. All responded that completing one request takes an average of two minutes. For the CTC2 database, all data clerks (100%) said the system was simple to use and requires an average of three minutes to complete one request. Since CTC2 identifies clients eligible for HIV viral load testing and case definitions were easy to understand, all data clerks at CTC reported that the process is simple. The data reporting structure, including data collection tools, was easy, and the case definition was easy to understand. The case definition included the timing of taking samples for testing at different intervals and the clinical condition status of the clients.

#### Flexibility

The HVL system’s electronic nature enables flexibility in system modifications, system addition, and data collection, allowing for continuous integration and updating of surveillance systems. The HVL surveillance system has adapted to the addition of the early infant diagnosis (EID) surveillance system, the HIV Testing Services (HTS) module, and the COVID-19 module were accommodated in the CTC2 database. The findings show that the system is flexible enough to add other surveillance systems.

#### Data Quality

Out of the five variables evaluated (age, gender, sample received date, results authorised date, and testing reason), an average of 97.4% of data was completed at the national level. Completeness ranged from 91.4% (result authorised date) to 100% (sample received data). The data correctness and consistency were evaluated at two facilities; 68% of the 41 laboratory findings selected and confirmed in the CTC2 database from DRRH and MHC were concordant that means data were consistency from the two databases when compared. Other data were discordant due to transcription errors. The overall data correctness at the two facilities reviewed was 96.8% correct entry to the system. Other errors observed where the system inserted the default date for the sample receiving date when not entered (Table 2).

**Table 2.**
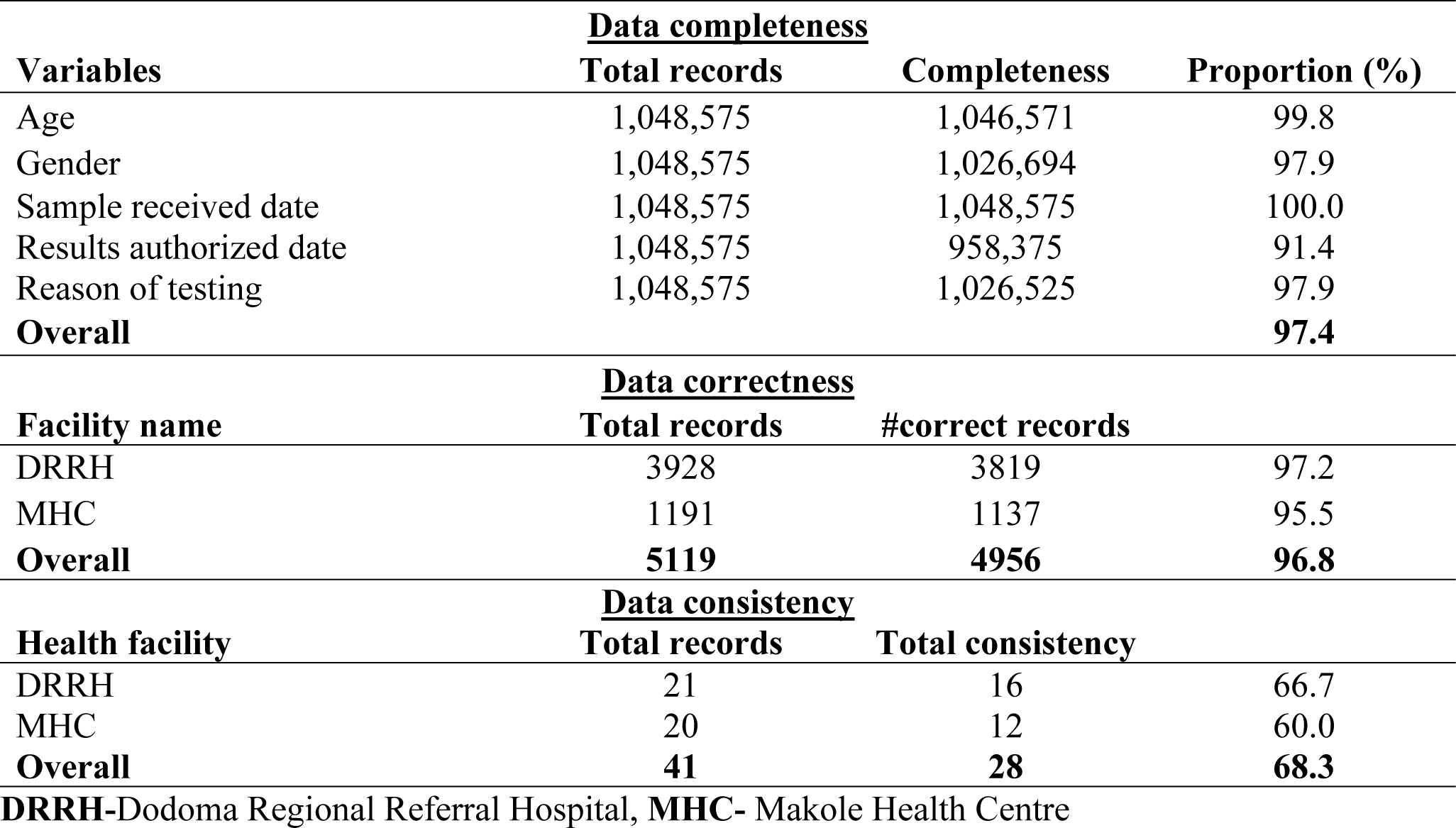
Viral load data completeness, correctness, and consistency at national and facility levels in the year 2022.

### Acceptability

All 8 (100%) data clerks interviewed had received training on the system and were willing to use it. The time it takes to process patient requests and submit information to the system shows users’ motivation regarding using it. The facilities that were collecting samples and participated in the system were 90.6% all over the country.

### Sensitivity and Positive value predictive

The proportion of the HVL tested from eligible clients was 84.4% (3134/3712) of all clients. The system detected 6019 eligible clients for HVL testing. Only 3134 of the 6019 eligible patients at the CTC2 had their samples collected between January and December 2022, making the system a Positive Value Positive of 52% (Fig 2).

**Fig 2.**
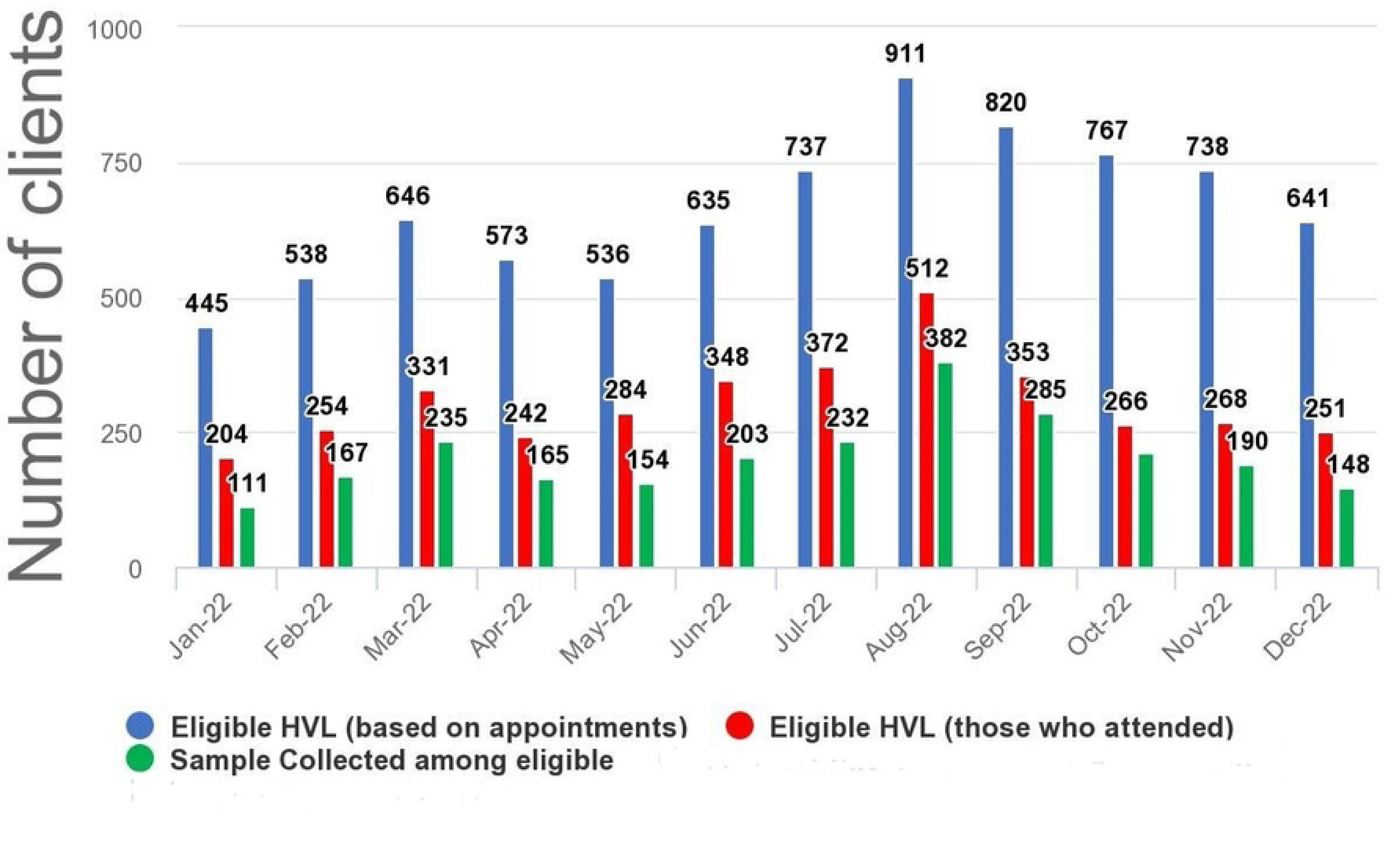
Graph showing the eligible clients based on appointment and sample collection status year 2022: Source CTC2 database: Dodoma Regional Referral Hospital.

### Representativeness

The HIV viral load surveillance system was representative in person, place, and time. Data was gathered and analysed nationwide from all hubs, spokes, and PCR testing laboratories. All regions were involved in the surveillance program. Persons’ demographic data was tracked, and socio-demographic information (e.g., age, gender) and clinical information, including laboratory test results, were collected. Over a period of one year, 2022, all known HIV populations were under surveillance. Over 90% of HVL-involved facilities sent samples to PCR testing laboratories nationwide (Fig 3).

**Fig 3.**
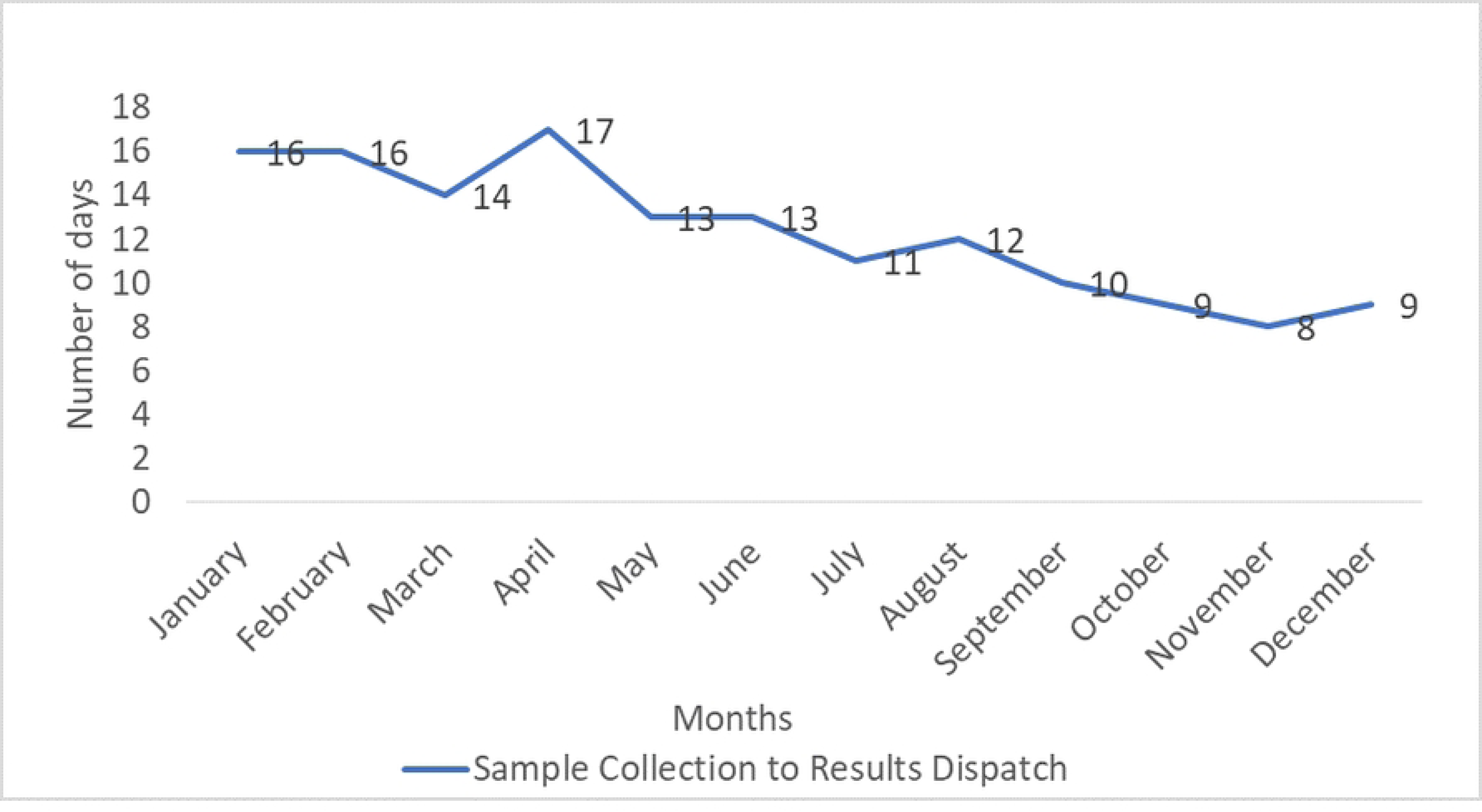
Map showing the distribution of HIV viral load sample-sending facilities in the year 2022.

### Timeliness

The system’s timeliness was assessed by one variable from the testing request generated from the spokes to the results returned. The national overall TAT ranged from 8 to 17 days, averaging 12 days (Fig 4). During data verification at the facility level, we found that the mean and median TAT were 14 and 13 days, respectively. During data evaluation, we found that 98.1% of samples were within the established national TAT of 14 days at DRRH and MHC (Table 3).

**Fig 4.**
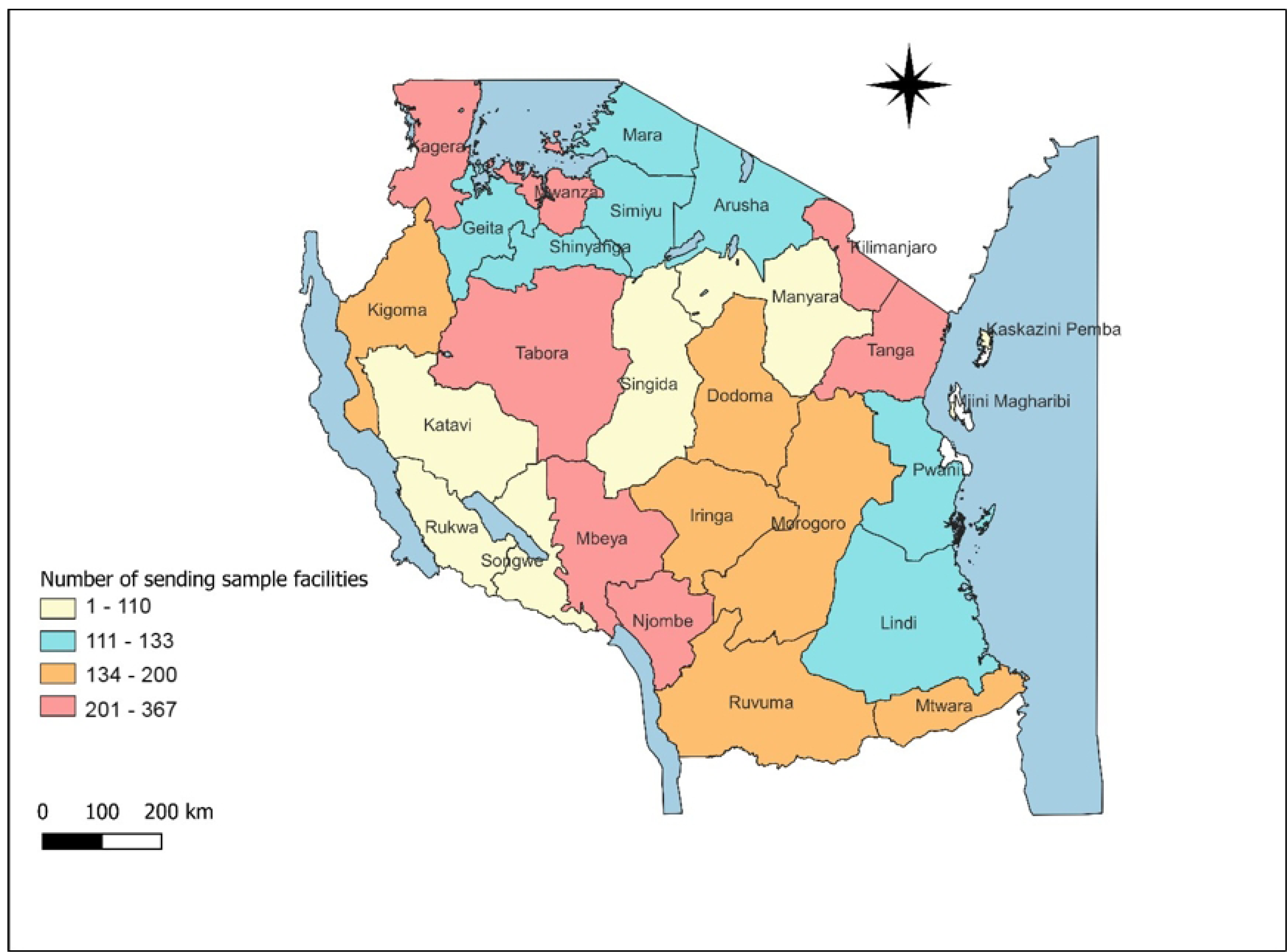
National HVL monthly TAT from sample collection to dispatch of result. Source: https://evlims.nacp.go.tz/#/

**Table 3:**
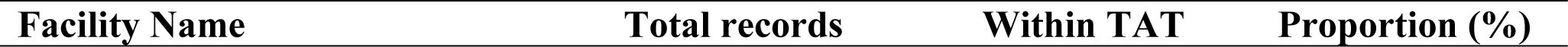
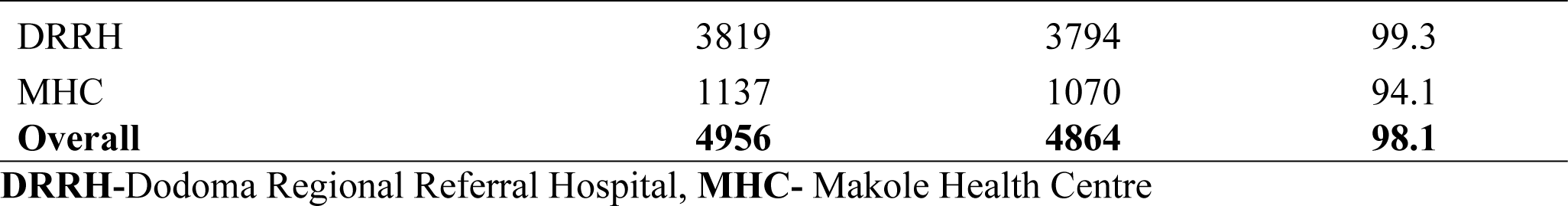
The number of records of TAT reviewed and verified per facilities, 2022.

### System stability

The CTC and laboratory information systems were stable quarterly for 98% and 95%, respectively, for the number of working days per quarter. Regardless of some challenges reported, it appears to have gone offline sometimes due to server problems and networking (Table 4)

**Table 4:** Number of average days reported for system functioning per one quarter, 2022.

| **System stability** | **Total time in use**<br>**(days)** | **Total time stable** | **Percentage (%)** |
| --- | --- | --- | --- |
| CTC 2 system | 120 | 117.6 | 98 |
| Laboratory Information system | 120 | 114.0 | 95 |
| **Overall system stability** | **240** | **231.6** | **96.5** |

## Discussion

The findings show that the HVL surveillance system was stable to an average of 96.5% working, sensitive by 84.4%, and representable to 90.6% of all facilities involved. The system was easy to use, and the turnaround time from sample collection to results was 98.1% verified at facilities. The system is reported to be useful for data reporting, visualisation, and early alerts provided by the system to make immediate program decisions. In people living with HIV, routine viral load testing enables the timely detection of suboptimal ART adherence and the proper management of treatment failure (9).

We found the national data completeness of the HVL surveillance system to be 97.4%, which was higher than the evaluation done in Atlanta and California, which shows an overall estimated completeness of reporting of 76% and 87%, respectively (10,11). Our findings’ high percentage of data completeness could be attributed to the system’s creation of a default date of 1970 for those requests without a sample receiving date, which increases the overall completeness. Furthermore, the data completeness could be attributed to close mentorship and supervision provided by the Ministry of Health to the spokes and hubs of HVL sites. The routine mentorship covers the area of data completeness and timely submission of data and samples to testing laboratories. However, despite the high percentage of data completeness, we found only 68% of the data were consistent, while 32% were discordant. The findings could be attributed to the manual entry of laboratory results from the laboratory information system to the CTC2 database. In addition, for clients who attended the CTC clinic at other centres, their data were not captured/entered into the sites due to a lack of data clerk accessibility of the clients’ accounts/electronic files.

We found that the surveillance system was representative of the coverage of 90.6% of all facilities in the country involved in the surveillance. The results were compared to a study done in South Africa, which shows the coverage of 98.3% (12). The high percentage of coverage could be attributed to country strategies of free HIV care and treatment services including consultation, testing, medications, and follow-up. We found a system sensitivity of 84.4%; this could be attributed to six multi-month dispensing (6MMD) in the country that delayed the eligible clients from taking HVL samples for testing, which started in 2022 (5). Regardless the challenge noted, it has been reported that giving stable customers multi-month dispensing (MMD) of ARV improves health outcomes in terms of immunosuppression, viral suppression, retention, mortality, and adherence (13–15).

For timeliness, we found that 75.6% of data reached the facility on time, from the testing laboratory to reporting back to the facility. Our findings report timeliness was higher than the study in South Africa, which found it to be 49.7% (12). The improvement observed was increased due to support provided by stakeholders including but not limited to financial support for mentorships and supervisions and technical assistance. The HVL data from the national perspective, including the facility’s continuous quality improvement, are discussed quarterly in the HIV Viral Load Technical Working Group (TWG), funded by the implementing partners.

We found that the TAT from sample collection to results authorisation was an average of 12 days, which could contribute to the increased timeliness of the report to 75.6% within the 14-day established target. This was high compared to Tanzania’s influenza sentinel surveillance, which shows a TAT of 36.6% within the target (16). Our findings of short TAT of 12 days could be attributed to the decentralisation of testing of HVL in the country and strong establishment of sample referral system.

The system can change under new operational conditions requiring less manpower, funds, or time. The system was adaptable to new health-related events, modifications to technology or case definitions, and changes in financing or reporting sources. Furthermore, the systems are easily interconnected with other systems and employ common data formats, such as those used in electronic data interchange. The system was flexible to accommodate new surveillance systems such as EID, HIV Testing Services (HTS), and COVID-19 vaccination surveillance for people living with HIV. The system modules added were interlinked to the main system, and the data flow uses the same business matrix. The system was interlinked with the electronic sample referral system, Tillelab, and other laboratory information systems. This was adaptive compared to the influenza sentinel surveillance system, which is still under manual data entry (16).

This study was subject to several limitations. It was a retrospective analysis, and we encountered that the system created a default date of 1970 for those requests without a sample receiving date, which made 3% incorrect receiving date and incorrect TAT. The large sample size used makes the study findings representable and generalizable.

## Conclusion

Overall, the system performs satisfactorily: It was acceptable and satisfied that it could monitor viral load suppression. Based on the system’s information, there will be a growing need to enhance the data and make it more usable. The absence of compatibility between the CTC2 database and laboratory information systems was the cause of the observed data discrepancy.

## Funding

The system evaluation was not funded.

## Author’s contribution

**PRT-** Conceptualization, Data curation, Formal analysis, Methodology and Writing – original draft, Writing – review & editing, **LJU** Conceptualization, Supervision and Writing – review & editing, **AM**- Methodology and Writing – review & editing, **ASM**- Data curation, Supervision and Writing – review & editing. **JNA**- Data curation, Writing – original draft and Writing – review & editing, **DO and JM**- Writing – review & editing, **FS and YB**- Data curation, **MVM** and **AJ**- Methodology, Supervision and Writing – review & editing

## Data Availability

All data produced in the present study are available upon reasonable request to the authors

## Acknowledgement.

We are very grateful to the Muhimbili University of Health and Allied Sciences, Ministry of Health, National AIDS Control Program (NACP), Tanzania Field Epidemiology and Laboratory Training program for technical support of this study

## Notes

### Competing Interest Statement

The authors have declared no competing interest.

### Funding Statement

This study did not receive any funding

### Author Declarations

Ethics committee/IRB of Ministry of Health gave ethical approval for this work Ref. no. PA.160/291/01A/41

